# Modelling the evolution of COVID-19 in high-incidence European countries and regions: estimated number of infections and impact of past and future intervention measures

**DOI:** 10.1101/2020.05.09.20096735

**Authors:** Juan Fernández-Recio

## Abstract

A previously developed mechanistic model of COVID-19 transmission has been adapted and applied here to study the evolution of the disease and the effect of intervention measures in some European countries and territories where the disease had major impact. A clear impact of the major intervention measures on the reproduction number (*R*_t_) has been found in all studied countries and territories, as already suggested by the drop in the number of deaths over time. Interestingly, the impact of such major intervention measures seems to be the same in most of these countries. The model has also provided realistic estimates of the total number of infections, active cases and future outcome. While the predictive capabilities of the model are much more uncertain before the peak of the outbreak, we could still reliably predict the evolution of the disease after a major intervention by assuming the afterwards reproduction number from current study. More challenging is to foresee the long-term impact of softer intervention measures, but this model can estimate the outcome of different scenarios and help planning changes in the implementation of control measures in a given country or region.

## INTRODUCTION

The ongoing pandemic expansion of the new SARS-CoV-2 virus has caused over 3,500,000 detected cases of coronavirus disease 2019 (COVID-19) and claimed over 240,000,000 lives worldwide as of 5 May 2020 [1]. After the first epidemic wave has dramatically hit a large number of countries, we can learn from all available epidemiological data to understand the evolution of the disease in order to better prepare the healthcare capacities for future pandemic waves. A recent model of SARS-CoV-2 transmission based on estimates of seasonality, immunity and cross-immunity from past virus pandemic data, indicates the high risk of recurrent outbreaks for the coming years, according to different scenarios [2]. Before the availability of effective vaccines and pharmaceutical treatments, the transmission of the disease can only be controlled by prevention measures, like personal hygiene habits, social distancing, case detection and isolation, or contact tracing. Strict social distancing enforced by the authorities has been the strategy of choice by the majority of countries in this first epidemic outbreak, implemented as different measures in each country (imposing or encouraging home confinement, closing schools and workplaces, or banning assemblies and gatherings, among others). Given that this usually involves a strong economical and behavioural impact in the society, it is urgently necessary to understand the evolution of the disease in each country or territory and estimate the effect of implementing such control measures. A major problem is that in most countries, only a small portion of the infected individuals is detected, and as a consequence, undetected contagious patients are the major cause of the expansion of the disease, which is the main limitation to foresee the effect of intervention measures. In the absence of large-scale detection tests applied to the entire country population, mathematical modeling can help to estimate the dynamics of infections based on available epidemiological data. Indeed, a recent networked dynamic metapopulation model using Bayesian inference and based on reported infections and mobility data suggests a high proportion of undocumented infections in the first SARS-CoV-2 outbreak within China [3]. That model compared the initial transmission dynamics with that after greater control measures were taken, showing a clear reduction in the transmission rate and the effective reproduction number. However, the specific impact of the different control measures was difficult to analyze at that time.

In this context, a Bayesian mechanistic model was recently proposed to estimate the effects of specific intervention measures on the reproduction number in 11 countries by inferring the number of infections from the observed deaths over time [4], assuming an infection fatality ratio (IFR) different from each country, adjusted to the age distribution of their population. A major assumption of that study was that the intervention measures had the same relative effect in all countries, which is not necessary true. For instance, according to Location History data from Google [5], the effect of lockdown measures on the mobility of the population was clearly different in each country. The level of isolation of transmitting cases not only depends on the intervention to the general population, but also on the percentage of detected patients, which might make more or less effective the isolation of such patients.

Another limitation of the mentioned study [4] is that, at that moment, the pandemia was at the early stages of the evolution. Since the peak of the outbreak had not yet been reached in the studied countries, the impact of intervention could be followed based solely on small changes in the slope of the growth of reported deaths over time, with a large degree of uncertainty. Moreover, for some of the intervention measures, not sufficient time had passed to see a real impact on the number of deaths. As a consequence, the impact of the intervention measures of most of the countries were probably underestimated, and the forecast anticipated a larger number of expected infections and deaths than that found in the following weeks.

Here, a similar study has been performed by using current data on the five European countries with the highest number of total cases (Spain, Italy, UK, France, and Germany), in which the disease has significantly evolved and seem to have overpassed the outbreak peak. In this work, the analysis has also included Iceland and the Spanish region of La Rioja, with comparable populations, in which the number of reported cases in proportion to their population is among the highest ones in Europe. The model has been adapted by fitting it individually to each country, and by simplifying the number of intervention measures to evaluate. The result is a model that nicely explains the evolution of the disease and that can describe the effective impact of the intervention measures and estimate the outcome future interventions.

## METHODS

### Data collection

Daily real-time death data was gathered from the ECDC (European Centre of Disease Control). Table 1 shows the up-to-date (5 May 2020) detected cases and reported deaths for high-incidence countries (Spain, Italy, United Kingdom, Germany, France, Iceland), as well as for La Rioja, the region in Spain with the highest-incidence relative to its population. In the latter, we have also shown the data after excluding the population from elderly retirement homes with reported cases. The high incidence of cases in elderly retirement homes in European countries suggests that transmission of disease has followed different patterns in these cases, and the impact of general lockdown measures might not be the same as in the general population. Since this data is available for this work in the case of La Rioja, it has been considered worthy to include it in the analysis.

**Table 1.** Reported COVID-19 cases per country/region (updated 5 May 2020)

| Country / Region | Total detected cases | Cases per 100K people | Total reported deaths |
| --- | --- | --- | --- |
| Spain | 219,329 | 469 | 25,613 |
| Italy | 211,938 | 351 | 29,079 |
| UK | 190,584 | 287 | 28,734 |
| Germany | 163,860 | 198 | 6,831 |
| France | 131,863 | 197 | 25,201 |
| La Rioja (Spain) | 3,969 | 1,256 | 336 |
| La Rioja (Spain)* | 2,988 | 952 | 143 |
| Iceland | 1,799 | 509 | 10 |

\*Excluding cases in elderly retirement homes. Reported deaths correspond to May 5<sup>th</sup>, 2020, but detected cases have been extrapolated from data on May 15<sup>th</sup>, 2020

### Estimating new infections and deaths over time

The major details of the model have been described elsewhere [4]. Basically, the expected number of deaths *D_i_* in a given day *i* is a function of the number of infections *I_j_* occurring in the previous *j=1…i-1* days, according to a previously calculated infection-to-death (*ITD*) probability distribution and an estimated infection fatality ratio (IFR) for each country (eq. 1).

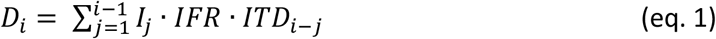

The infection-to-death (*ITD*) probability distribution was modelled, using previous epidemiological data [6,7], by adding up two independent distributions: i) the infection-to-onset (incubation period) distribution, estimated as a Gamma function with mean 5.1 days and coefficient of variation 0.86, and ii) the onset-to-death (time between onset and death) distribution, estimated as a Gamma function with a mean of 18.8 days and a coefficient of variation 0.45 [7]. The resulting *ITD* probability distribution is shown in Figure 1B.

**Figure 1.**
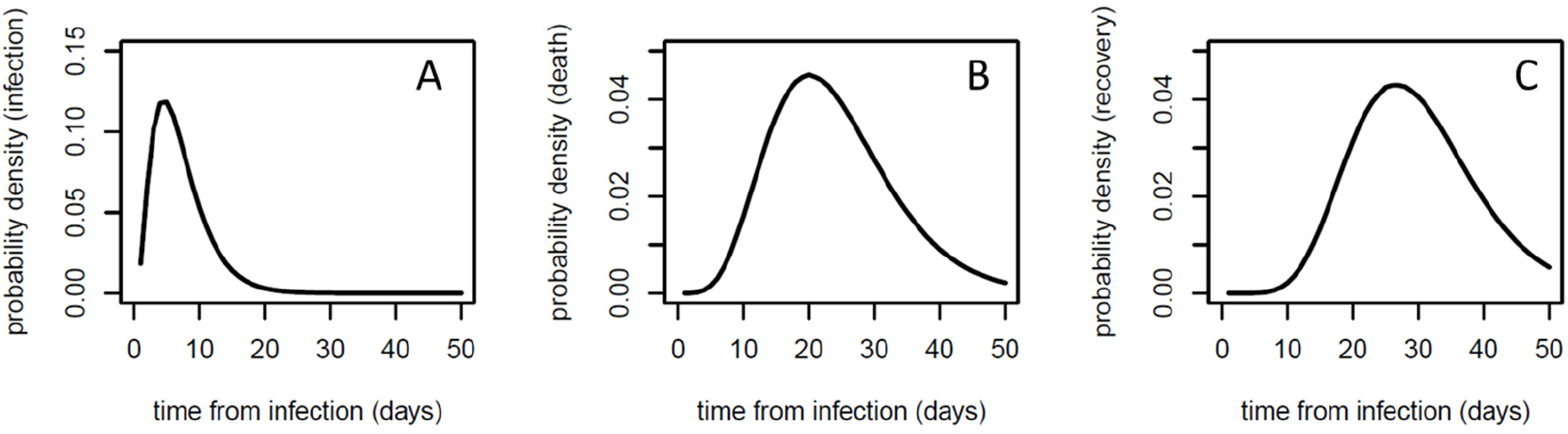
Serial interval (A), infection-to-death (B), and infection-to-recovery (C) probability distributions used in this work.

The infection fatality ratio (IFR) is the probability of death for an infected case. The values used here for COVID-19 are derived from previous estimates [8] and are calculated for each country according to the age-distribution population [6,7]. For La Rioja, the same IFR was used as for Spain. For Iceland, the general IFR value of 0.657% was initially used, as previously reported by combining estimates of case fatality ratios with information on infection prevalence in China [7]. This IFR estimate provided an expected number of infected and active cases close to the detected ones. However, the case fatality rate (CFR), an empirical value obtained from the number of deaths over the detected cases, gives a value of CFR=0.556% for Iceland, and the true IFR could be even smaller [9], so this CFR value (0.556%) was finally used as a better estimate for IFR in Iceland. The IFR values used in the study here for the different countries are shown in Table 2.

**Table 2.** Model parameters (intervention dates and IFR) used in this work

| Country / region | Time of intervention | Days from first 100 cases to intervention time | IFR |
| --- | --- | --- | --- |
| Spain | 14 March | 11 | 0.926% |
| Italy | 11 March | 16 | 1.090% |
| UK | 24 March | 18 | 0.919% |
| Germany | 22 March | 20 | 1.093% |
| France | 17 March | 15 | 1.153% |
| La Rioja | 14 March | 5 | 0.926% |
| Iceland | 24 March | 8 | 0.556% |

In this model, the expected number of new infections *I_j_* occurring in a given day *j* is a function of the number of infections *I_k_* in the previous *k=1…j-1* days, according to a serial interval (SI) distribution, and the reproduction number (*R_t_*) (eq. 2).

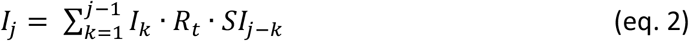

The *SI* probability distribution was modelled as a Gamma function with a mean of 6.5 days and a coefficient of variation 0.62 (eq. 2), as previously described [4], and is shown in Figure 1A. The reproduction number (*R_t_*) had an initial fix value *R_0_* (a parameter that was left to optimize during fitting of the model independently for each country), and was assumed to change after an intervention to another value which is kept fix over time until a new intervention (the new *R_t_* value after a given intervention is also a parameter to optimize during the model fitting).

### Model fitting

The expected number of deaths obtained from the above described model (eq. 1 and 2) were fitted to the observed number of daily deaths, assuming to follow a negative binomial distribution, as previously described [4]. In the case of Iceland, both the observed number of daily deaths and the cumulative number of observed deaths were fitted, because the observed daily deaths alone did not provide reliable fitting due to small size of sample. Following the initial procedure [4], the model included only observed deaths from the day after a given country had cumulatively observed over 10 deaths (*d_10_*), given that the early stages of the epidemic in a given country might be dominated by infections that are not local. An exception was done in the case of small regions or countries with low number of deaths, such as Iceland and La Rioja, in which observed deaths were included from the day of the first death (*d_1_*). Similarly, according to the original procedure [4], the initial infections of the model were assumed to be 30 days before this *d_10_* (or *d_1_*) day, starting with 6 consecutive days with the same number of infections, which was left as a parameter to optimize in the fitting procedure. The initial infections for the first 6 days and the *R*_0_ and *R*_t_ values after each intervention were defined as parameters to optimize in the model. Fitting was done in the probabilistic programming language Stan, using an adaptive Hamiltonian Monte Carlo (HMC) sampler. Eight chains for 4000 iterations, with 2000 iterations of warmup and a thinning factor 4 were run. Running of 200 sampling iterations with 100 warmup iterations yielded very similar results in most of the cases, suggesting that convergence was achieved early in the fitting process. See more details in the original description of the model [4]. The original code is available at https://github.com/ImperialCollegeLondon/covid19model/releases/tag/v1.0

In the original study, parameters were estimated simultaneously for 11 countries, but the model was fitted to the data from each country/region independently, since the effect of each intervention on *R*_t_ is not necessarily the same in all societies. To simplify the model, here the number of interventions were reduced, and the possibility of having an end date for any intervention was added. The inclusion of only one intervention was sufficient to explain the data, while the addition of further intervention steps did not significantly improved the fitting.

### Predictive model from a given set of parameters

The use of continuous serial interval and infection-to-death probability distributions to estimate the number of deaths from the new infections is needed for the efficiency of the fitting procedure, but in reality, the deaths derived from the new infected people on a given day will be distributed in specific days in a discrete manner. The overall discrete distributions will expectedly follow the serial interval and infection-to-death probability distributions, but the number of deaths for each set of infections will happen in a single discrete distribution, different each time, especially if the number of infections is small. In Figure 2 we can compare some random discrete distributions for expected deaths over time for samples of different size (*n*=1, 10, 100, 1000) according to the continuous infection-to-death probability distributions used here vs. several discrete samples derived from the same infection-to-death distribution. As the sample size (*n*) increases, the discrete and continuous distributions tend to converge, but for small size samples, assuming continuous distribution might not represent a given instance.

**Figure 2.**
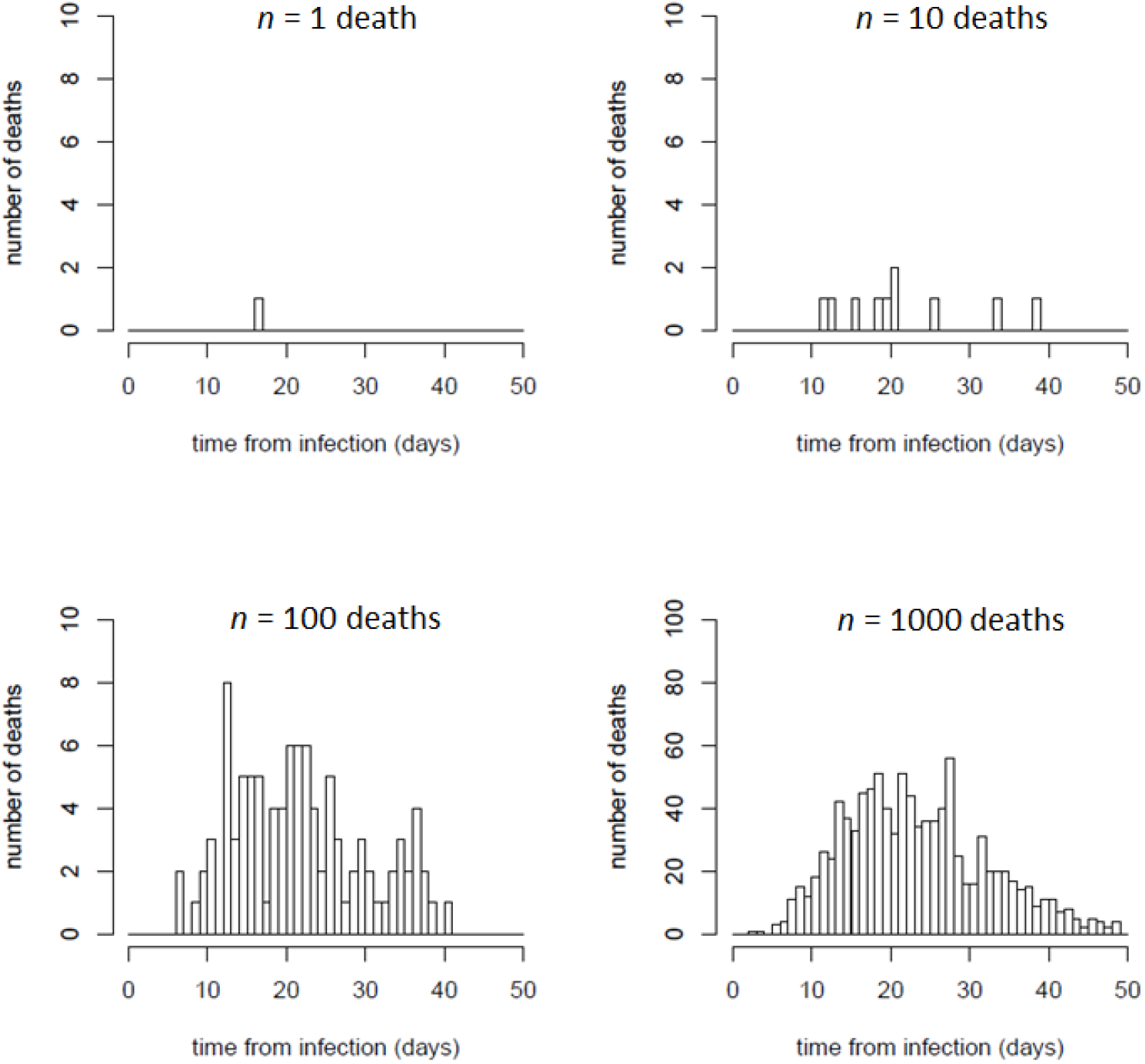
Examples of random discrete distributions of samples of *n*=1, 10, 100, 1000 expected deaths, according to the infection-to-death (ITD) probability distribution used in this work. For samples of small size, the distribution can be very different from the probability curve shown in Figure 1B, while for samples of larger size (e.g. 100 or 1000 deaths), the discrete distributions get closer to the probability distribution curve.

Therefore, for a better visual comparison between the predictive results of our model and the set of reported deaths over time (discrete distribution), it is possible to generate instances of discrete distributions over time of the expected number of new infections and deaths computed with the parameters obtained during fitting procedure (usually, from a set of parameters randomly selected among the ones obtained). These instances can be generated by randomly assigning every case (from the set of new infections or deaths to be distributed) to a specific day according to the serial interval and infection-to-death probability distributions.

### Estimating active cases from model predictions

Basically, the expected number of recovered cases *RECOV_i_* in a given day *i* is a function of the number of the estimated new infections *I_j_* occurring in the previous *j=1…i-1* days, according to a previously calculated infection-to-recovery (*ITR*) probability distribution, after discounting the percentage of new infected cases with outcome of death from infection fatality ratio (IFR) (eq. 3).

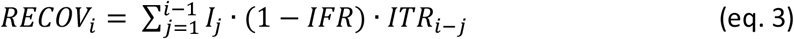

The infection-to-recovery (*ITR*) probability distribution was modelled by adding up two independent distributions: i) the infection-to-onset (incubation period) distribution, estimated as a Gamma function with mean 5.1 days and coefficient of variation 0.86, and ii) the onset-to-recovery (time between onset and recovery) distribution, estimated as a Gamma function with a mean of 24.7 days and a coefficient of variation 0.35 [7]. The resulting *ITR* probability distribution is shown in Figure 1C. The estimated active cases for a given day is calculated by subtracting the estimated cumulative number of recovered cases from the estimated cumulative number of cases for that day.

## RESULTS

### Model suggests a significant impact of intervention measures on disease transmission

The model was fitted (see Methods) to data from the countries with the highest number of reported cases in the EU/EEA and the UK as for May 5^th^, 2020 (Spain, Italy, United Kingdom, Germany and France) (Table 1). They are actually the countries with the largest population. For comparison, the model has also been applied to small countries and regions, such as Iceland, and the Spanish region La Rioja, which have comparable population (over 300K inhabitants) and show a high incidence of cases per 100K people. Actually, La Rioja is the region with highest number of detected cases relative to its population in Spain and probably in Europe. For comparison, the region in the United States with the highest-level of incidence is New York City, with boroughs like The Bronx with 2,667 detected cases per 100K as for 5 May 2020 [10].

It has been evaluated here the effect of the single most relevant intervention measure implemented by authorities (defined as “lockdown ordered” in a previous study [4]). For La Rioja, the same lockdown date was used as for Spain (March 14th). For Iceland, the date of 24 March has been considered, when a nation-wide ban was enforced on public assemblies over 20, as well as the closure of bars and most public businesses. The dates of these interventions are shown in Table 2. After fitting the model for each country/region independently, the estimated number of daily infections and deaths derived from the model are shown in Figure 3, in comparison with the reported ones. The resulting reproduction number values derived from the model provide an estimation of the impact of the intervention measures on the transmission dynamics of the disease in each country (Table 3).

**Figure 3.**
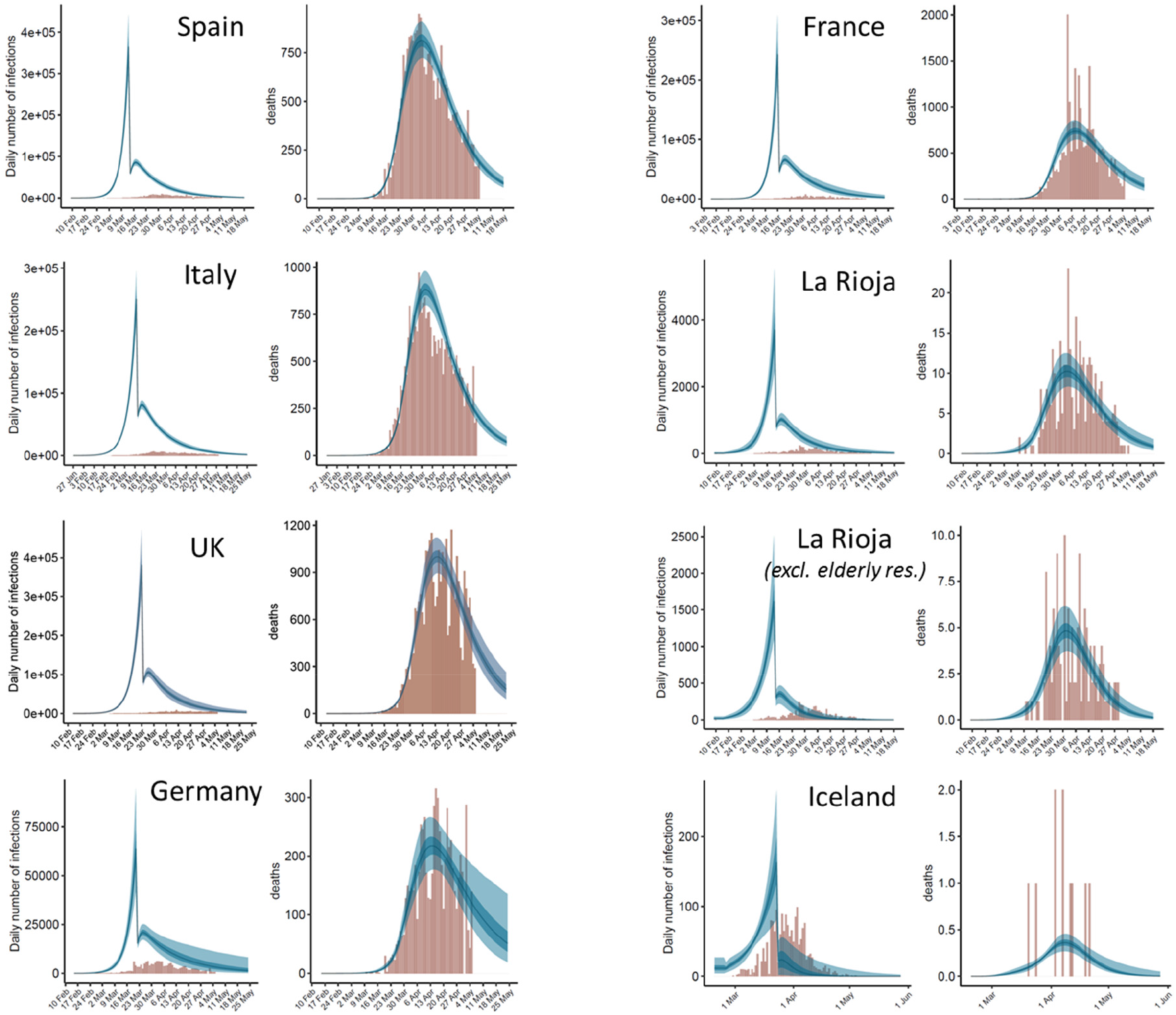
Estimates of infections and deaths over time after independent application of the model to each country and region. For each country, the left plot shows the predicted daily infections as compared with the reported ones, and the right plot shows the expected daily deaths as compared with the observed ones. Expected values are shown as blue bands: light blue (95% CI), dark blue (50% CI) and line (median). Observed values are shown as brown bars. The estimates of *R*_t_ before and after intervention resulting from the model are the ones in Table 3. In all cases, after intervention, *R*_t_ is significantly reduced to a value well below 1 and the number of new infections decreases. In La Rioja second panel, daily observed deaths do not include those from elderly retirement homes, but daily reported cases are the total ones, since the daily reported cases outside elderly residences was not available for this work.

We can observe that the final *R_t_* after major intervention is similar in all analyzed countries (except Iceland, and La Rioja when elderly residences with reported cases were excluded), with mean values ranging from 0.57 (La Rioja region) to 0.71 (Germany), and an averaged value of 0.625. Interestingly, a recent calculation on the reproduction number (R) in Germany, estimated from a nowcasting approach on reported COVID-19 cases with illness onset up to 3 days before data closure, provides a current estimate of R=0.71 (95% prediction interval: 0.59-0.82) [11], which is virtually the same as the one calculated here with the disease transmission model based on the reported deaths. Regarding the relative values of *R_t_* after intervention (in percentage relative to *R_0_* before intervention), they ranged from 12.0% (Spain) to 20.7% (Italy). These values seem to depend not only on the effectiveness of the intervention measures, but also on the evolution of the disease prior to intervention, described by *R_0_*, which seems to be different in each country (Table 3). As a warning note, this effect in relative terms was the one assumed to be fix for the same type of intervention in the different countries in the original application of the model [4], an assumption that does not seem to be valid.

**Table 3.**
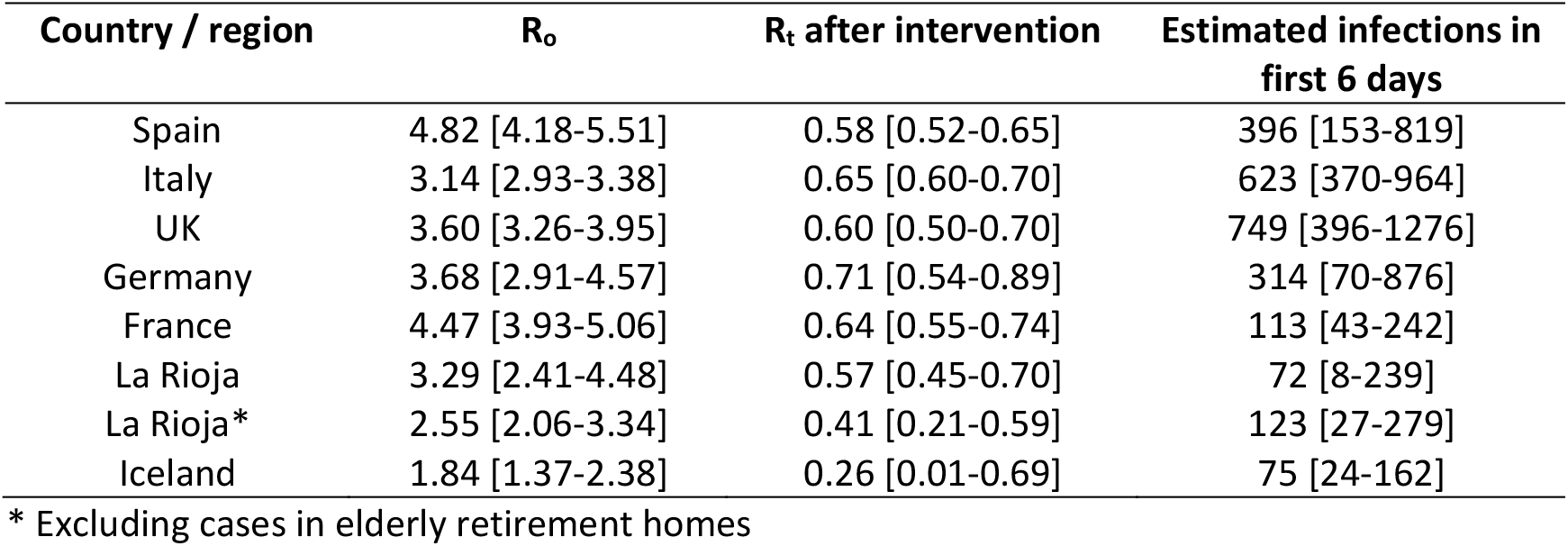
Estimated reproduction numbers (before and after intervention) and initial number of infections resulting from the model (shown mean values, with 95% credible interval)

The model has also been applied by including additional interventions, such as a work lockdown period ordered in Spain and La Rioja region between 1 and 6 April 2020 (see Supplemental Table 1), or a contention period around 7 March 2020 in La Rioja after a very localized outbreak in the city of Haro (data not shown), but no significant improvement of the model was found (Supplemental Figure 1). The effect of an additional indirect intervention in Iceland was also modelled (on April 3^rd^, a recently released mobile app to trace infections had almost 75,000 downloads, which might have had an impact in the reproduction number), with no significant changes (Supplemental Table 1, Supplemental Figure 1). In general, one would expect that a series of stepwise interventions will have a smoother effect on the reproduction number (and therefore in the number of new infections) that just a single strict intervention. One could also speculate whether other early measures taken in some countries have contributed to decrease their initial *R*_0_ values or to delay initial number of infections at the early stage of the epidemic outbreak, but this is beyond this study.

The model has also been applied to La Rioja data after excluding data in elderly retirement homes with reported cases (Figure 3, Table 3). As expected, the propagation of the disease in the elderly retirement homes and in the general population is very different. Outside these elderly retirement homes, the propagation rates decrease, before and after intervention. It would be interesting to know whether the same effect can be seen in the rest of countries.

### Estimated number of total infections and active cases

From the values obtained by the model, we can estimate the daily cumulative number of infections. The evolution of the predicted cumulative number of infections in comparison with the detected cases over time can be seen in Figure 4, with the detailed data per country as for May 5^th^, 2020 in Table 4. The highest detection rates (that is, total detected cases with respect to the total predicted ones) are found in Germany (21%) and Iceland (54%). These are actually countries with a high number of tests performed relative to their population. In Germany, there were reported 30.4 tests per thousand people as for April 22, 2020 [12], one of the highest rates in Europe. In Iceland there were reported 151.2 tests per thousand people as for May 5^th^, 2020, the highest rate in Europe [13]. In the rest of the territories, the total number of predicted cases is much higher than the reported ones, with detection rates that range from 5% (UK) to 10% (La Rioja). Interestingly, the detection rate in La Rioja is better than in Spain (7%). Moreover, when data from elderly retirement homes with reported cases are excluded, the detection rate in the rest of population in La Rioja is much larger (18%). This is consistent with the fact that La Rioja was the Spanish region with the highest number of PCR tests per thousand people (56.9, as for Apr 27, 2020; as compared to 22.0 in Spain) [14].

From the model results, we can also estimate the number of daily active cases per country (Figure 4). For comparison, the reported number of active cases in La Rioja [15] and in Iceland [13] are shown. In La Rioja, there were reported 1,257 active cases as for May 5^th^, 2020. As indicated in Table 4, the model estimated a median value of 4,746 for the same date (95% CI: 2,821-8,088), which indicates a detection rate of 27%. When data from elderly retirement homes were excluded, the model estimated a median value of 945 active cases (95% CI: 464-2,059). Considering that the reported 1,257 active case as for May 5^th^ include cases from elderly residences (e.g. 112 out of the 741 active cases as for May 15^th^ were from elderly retirement homes [15]), this indicates a detection rate of active cases close to 100%. In Iceland, there were reported 32 active cases as for May 5, 2020, while our model predicted a median value of 141 active cases for the same date (95% CI: 78-605) (Table 4), indicating a detection rate of 23% (which is lower than the general detection rate for the total cumulative cases).

**Table 4.** Estimated cumulative infections as for 5 May 2020 (shown median values with 95% credible interval)

| Country / region | Estimated total infections | Detection rate | % population infected | Estimated active cases |
| --- | --- | --- | --- | --- |
| Spain | 2,990K [2,742K-3,269K] | 7.3% [6.7-8.0%] | 6.4% [5.9-7.0%] | 415K [329K-538K] |
| Italy | 3,094K [2,868K-3,358K] | 6.9% [6.3-7.4%] | 5.1% [4.7-5.6%] | 445K [365K-551K] |
| UK | 3,800K [3,406K-4,292K] | 5.0% [4.4-5.6%] | 5.6% [5.0-6.3%] | 998K [739K-1,369K] |
| Germany | 793K [641K-1,024K] | 20.6% [16.0-25.6%] | 0.9% [0.8-1.2%] | 245K [142K-451K] |
| France | 2,351K [2,089K-2,693K] | 5.6% [4.9-6.3%] | 3.6% [3.2-4.1%] | 482K [341K-715K] |
| La Rioja | 38,505 [32,850-45,155] | 10.3% [8.8-12.1%] | 12.2% [10.4-14.3%] | 4,746 [2,821-8,088] |
| La Rioja* | 16,205 [13,095-19,972] | 18.4% [15.0-22.8%] | 5.2% [4.2-6.4%] | 945 [464-2,059] |
| Iceland | 2,029 [1,669-2,647] | 88.7% [68.0-100%] | 0.6% [0.5-0.7%] | 141 [78-605] |

\* Excluding data from elderly retirement homes

**Figure 4.**
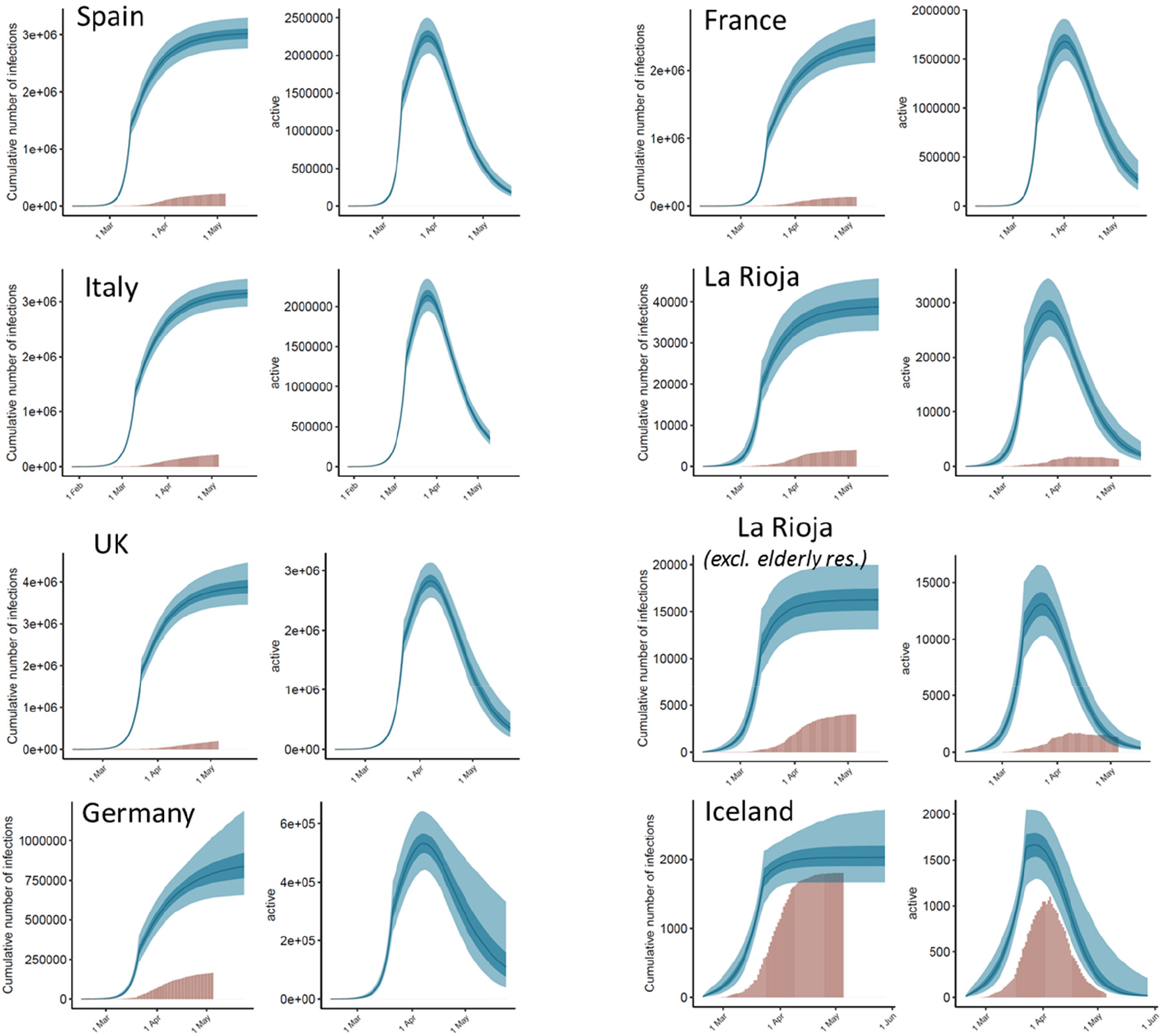
Estimated cumulative infections and active cases from independent application of the model to each country and region. For each country, the left plot shows the estimated cumulative infections for every day as compared with the reported ones, and the right plot shows the expected active cases each day (in some countries, they are compared with the reported ones). Expected values are shown as blue bands: light blue (95% CI), dark blue (50% CI) and line (median). Observed values are shown as brown bars. The estimates of *R*_t_ before and after intervention resulting from the model are the ones in Table 3. In all cases, after intervention, *R*_t_ is significantly reduced to a value well below 1 and the number of new infections decreases. In La Rioja second panel, estimates are derived from observed deaths after excluding those from elderly retirement homes, but accumulated infections and active cases over time are the total ones, since the daily cases excluding elderly retirement homes was not available for this work.

### Predicting discrete distributions of infections and deaths

A continuous distribution of the expected new infected cases and deaths provides a smooth curve that is suitable for parameter optimization. However, in reality, the infections and deaths are obviously distributed in a discrete manner. For better visualization of the predicted data, I have randomly generated discrete distributions of the expected infections and deaths, according to the continuous probability distributions and the parameters resulting from the model (see Methods). Figure 5 shows some discrete samples of the predicted infections and deaths, which resemble better the roughed distribution of the reported data, especially in small countries and regions.

**Figure 5.**
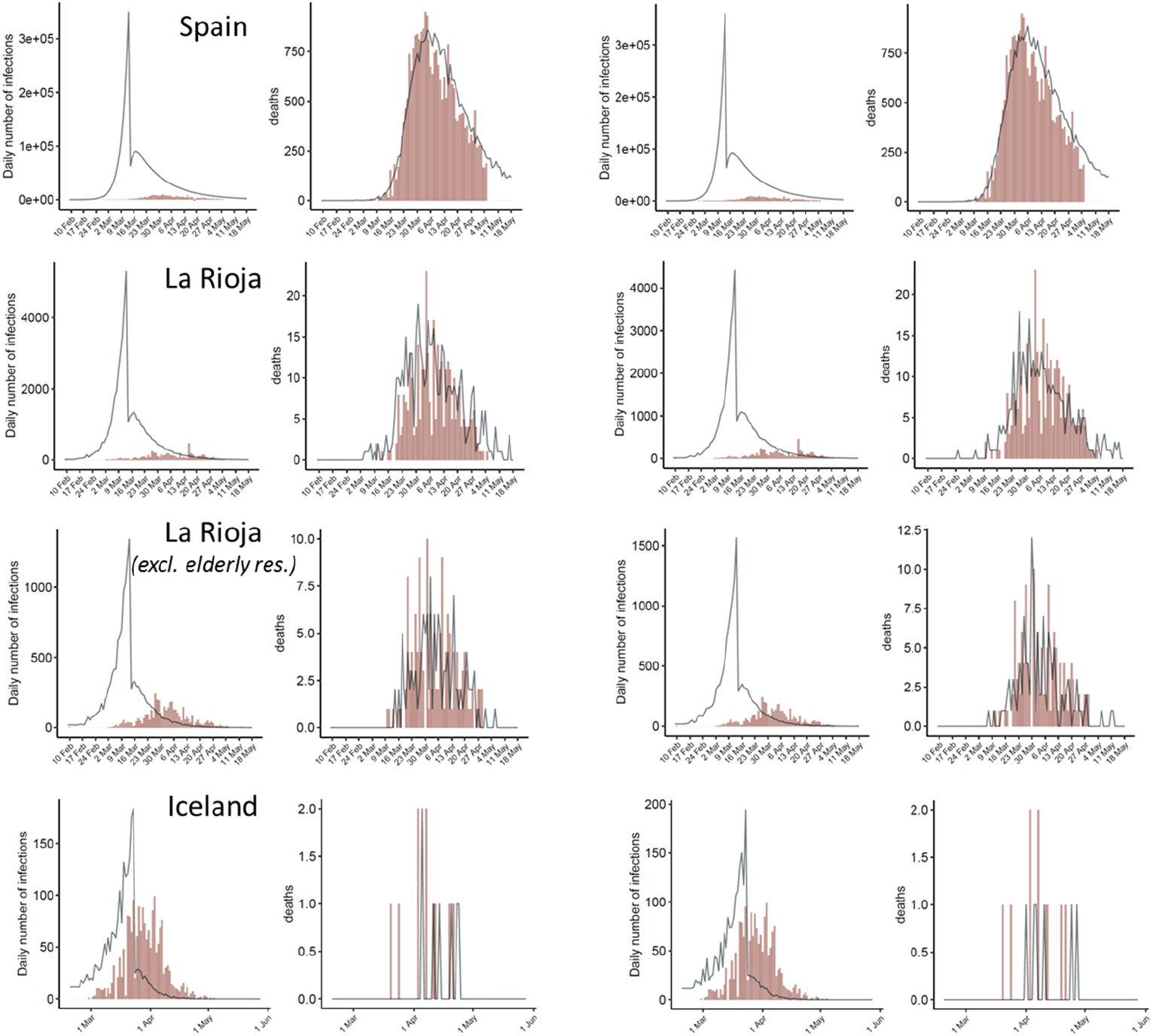
Instances of discrete distributions of the estimated number of daily infections and deaths randomly sampled according to the distribution probabilities of serial interval and infection-to-death, using the sets of reproduction numbers and initial infections obtained by the fitted model. When number of daily infections and deaths is large (e.g. Spain), the predicted discrete distributions are closer to the continuous ones (Figure 3), but when these numbers are smaller (e.g. La Rioja and Iceland), the discrete distributions resemble better the rough distribution of reported data over time. In La Rioja second panel, observed deaths do not include those from elderly retirement homes, but daily reported cases are the total ones, since daily cases excluding elderly retirement homes was not available for this work.

## DISCUSSION

### The reliability of the predictions depends on the stage of the epidemic outbreak

The credible intervals of the parameters and estimated data provided by the model indicate the reliability of the predictions. In the graphical plots of estimated daily cases and deaths (Figure 3) we can visualize the predictions for a few days beyond the last day of data (May 5, 2020). In general, in countries where more time has passed from the peak of the outbreak (e.g. Spain, Italy), the variability in the estimated data beyond the last day of data is smaller than in countries in which fewer days have passed from the time of the peak (e.g. Germany, UK). As a validation test, in order to evaluate the capabilities of the model to predict the evolution of the disease beyond current date, it has been fitted again to Spain and La Rioja data (with and without elderly residences cases), after removing the reported deaths corresponding to the last week, the two last weeks or the three last weeks of data in Spain and La Rioja (Figure 6B-D,H-J,N-P).

**Figure 6.**
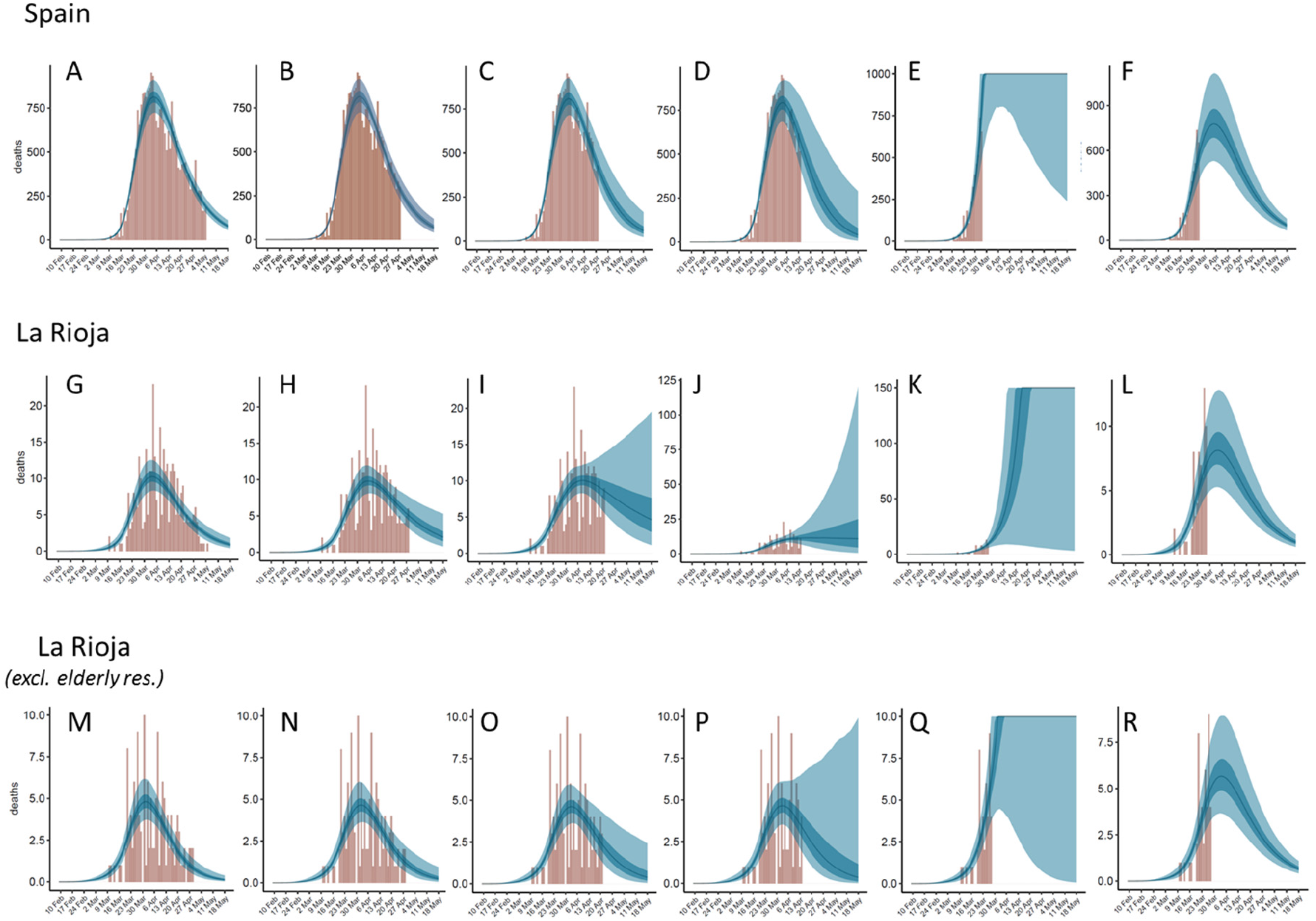
Estimated deaths for Spain (upper row) and La Rioja (total data: middle row; excluding elderly residences: bottom row) derived from the model (A,G,M) in comparison to the predictions obtained after removing last week (B,H,N), 2 last weeks (C,I,O), 3 last weeks of data (D,J,P), or keeping data just up to 1 week to the peak (E,K,Q), the latter also after keeping fix *R*_t_=0.625 (F,L,R).

Table 5 shows the reproduction numbers, and the estimated number of deaths for the last week of data (April 29 to May 5) obtained by the model when different sets of dates are removed. In general, we can observe that the more dates are removed and the fewer days left after the peak of the outbreak, the more uncertain the predictions get. Indeed, the predicted *R_t_* values obtained after removing larger sets of dates increasingly deviate from the ones obtained with the entire set of dates. Similarly, the number of deaths predicted for the last week of data increasingly deviate from the reported ones when larger sets of dates are removed. However, the response of the model to removal of data is different in the two cases analyzed here. In the case of Spain, the mean vale for the predicted deaths in the last week does not dramatically deviate from the real one even when 3 weeks are removed, although the uncertainty increases (the 95% CI range increases with respect to the ones obtained with the entire set of dates). We could safely say that three weeks ago the model would have been able to reasonably predict today’s situation in Spain (as for May 5^th^, 2020). In the case of La Rioja, the predictions get worse much faster upon removal of data, with larger deviations of *R_t_* and worse prediction of deaths for the last week of reported data. Perhaps this different behaviour of the model with shorter data is related to specific features of the disease evolution in each territory, or maybe it is simply due to differences in sample size (i.e. removing dates from regions with already small number of reported cases can introduce larger uncertainty).

**Table 5.** Reproduction number and expected deaths during the last week of reported data (29Apr-5May) after removing different sets of dates from the model

| Country / region | Forecast start time | $R_0$ | $R_t$ | Deaths 29Apr-5May |
| --- | --- | --- | --- | --- |
| *Spain* |  |  |  | 1791 (real) |
|  | 5 May (last day, original model) | 4.82 [4.18-5.51] | 0.58 [0.52-0.65] | 1653 [1386-1980] |
|  | 28 Apr (1 week to last) | 4.87 [4.20-5.63] | 0.57 [0.48-0.65] | 1562 [1178-2048] |
|  | 21 Apr (2 weeks to last) | 4.88 [4.13-5.69] | 0.54 [0.40-0.70] | 1470 [853-2466] |
|  | 14 Apr (3 weeks to last) | 4.90 [4.03-5.84] | 0.50 [0.23-0.78] | 1344 [430-3435] |
|  | 27 Mar (1 week to peak) | 4.08 [3.28-5.06] | 2.99 [0.78-4.37] | off-limits [2972-off] |
| | 27 Mar (1 week to peak) fix $R_t$ | 4.56 [3.57-5.66] | 0.625 | 1864 [1223-2666] |
| *La Rioja* |  |  |  | 10 (real) |
|  | 5 May (last day, original model) | 3.29 [2.41-4.48] | 0.57 [0.45-0.70] | 19 [13-28] |
|  | 28 Apr (1 week to last) | 3.00 [2.33-4.17] | 0.71 [0.55-0.88] | 31 [18-50] |
|  | 21 Apr (2 weeks to last) | 2.81 [2.24-3.83] | 0.84 [0.61-1.10] | 52 [22-111] |
|  | 14 Apr (3 weeks to last) | 2.72 [2.17-3.74] | 0.98 [0.54-1.44] | 108 [16-358] |
|  | 28 Mar (1 week to peak) | 2.76 [2.22-3.69] | 2.29 [0.80-3.35] | off-limits [37-off] |
| | 28 Mar (1 week to peak) fix $R_t$ | 2.86 [2.22-4.00] | 0.625 | 19 [11-29] |
| *La Rioja (excluding elderly residences)* |  |  |  | 2 (real) |
|  | 5 May (last day, original model) | 2.55 [2.06-3.34] | 0.41 [0.21-0.59] | 5 [2-8] |
|  | 28 Apr (1 week to last) | 2.46 [1.97-3.15] | 0.51 [0.29-0.72] | 7 [3-13] |
|  | 21 Apr (2 weeks to last) | 2.45 [1.93-3.14] | 0.58 [0.28-0.87] | 10 [3-24] |
|  | 14 Apr (3 weeks to last) | 2.44 [1.87-3.12] | 0.58 [0.14-1.10] | 13 [2-57] |
|  | 28 Mar (1 week to peak) | 2.53 [2.04-3.34] | 1.88 [0.32-2.88] | off-limits [3-off] |
| | 28 Mar (1 week to peak) fix $R_t$ | 2.61 [2.06-3.59] | 0.625 | 13 [8-20] |

It is interesting to evaluate the predictive capabilities of the model when using data just up to 1 week before the peak of the outbreak. In this case (Figure 6E,K,Q), predictions are much more uncertain and completely wrong. As we can see in Table 5, while the *R_0_* values are not dramatically different, the resulting *R_t_* values are completely wrong, which indicates that the model cannot estimate the impact of the intervention on the reproduction number before achieving the peak of the outbreak. This was actually the situation of the original study on 11 countries on March 28 [4], when the peak of the outbreak was still >1 week away in the majority of the analyzed countries. Consistent with the validation test here, in those conditions (>1 week before the peak) the model underestimated the impact of the different intervention measures and predicted a much higher number of infections and death than the reported ones in the upcoming days.

Fortunately, as above mentioned (see Results), the values obtained for *R_t_* (after a major intervention) seem to be quite consistent among all countries (except Iceland), with mean values between 0.57 and 0.71, and an averaged value of 0.625. Therefore, the model was fitted again to the short set of data (up to 1 week before the peak), but this time assuming a fix value of *R_t_* = 0.625. Remarkably, in these conditions, the predictions are actually quite good (Figure 6F,L,R), indeed comparable to those obtained with the entire set of data. This validation test suggests is that in cases in which the epidemic outbreak is not clearly over the peak, especially when the available data is noisy (e.g. small sample size), it could be a better option to apply the model using a guess value of *R_t_*=0.625 rather than trying to predict such *R_t_* value by fitting.

### Modelling long-term disease progression in different scenarios

The model above described is a useful tool to evaluate the evolution of the disease in each country or community. In these moments, it is urgent to evaluate the possible outcome of the planned changes in current transmission control measures. For instance, in Spain (including La Rioja), there are three phases proposed for the gradual return to normality.

Two key dates are 11 May 2020, the beginning of Phase 1 for most of the regions, in which meetings of up to 10 people will be allowed, and bar terraces will be opened (with some limitations), and 22 June 2020, the proposed date for the end of Phase 3, in which some regions can go back to a certain “normality”, allowing sports and cultural shows (with some conditions), as well as national travel. We can devise different scenarios regarding the potential effect on the reproduction number due to the gradual removal of lockdown measures in these key dates. Some options are: i) no changes in *R_t_* (very unlikely), ii) slight increment to *R_t_* =0.71 similar to current situation in Germany (also quite unlikely, given the activities that are planned to be allowed), iii) further increment to *R_t_* = 1.0, which implies doubling the number of patients actively transmitting the disease (this could be a likely scenario for the phase between 11 May and 22 June), or iv) much larger increment to *R_t_* = 1.8, similar to the situation in Iceland at the beginning of the epidemic, e.g. normal activities allowed but with extensive testing and isolation of detected infections (this is a likely scenario after 22 June). All these options have been considered for Spain and La Rioja, and the results are shown in Supplemental Figure 2 (Spain), and Supplemental Figures 3,4 (La Rioja). There is a further possible scenario that we can evaluate, which is the possible return to total normality in 1 September 2020, with open schools, usual sports and cultural shows, etc., so this variable has also been considered in Supplemental Figures 5 (Spain) and Supplemental Figures 6,7 (La Rioja). We can see that in some of these scenarios, the possibility of a new epidemic outbreak is clear. Figure 7 shows in more detail the predicted evolution for Spain in La Rioja (with and without elderly retirement home data) in a likely scenario, with *R_t_* = 1.0 between 11 May and 22 June, and *R_t_* = 1.8 afterwards. Assuming these reproduction numbers, we can foresee an outbreak in September/October. Obviously, the model does not consider the possible intervention measures to limit the impact of this hypothetical outbreak, which might depend on how early the potential new infections could be detected.

**Figure 7.**
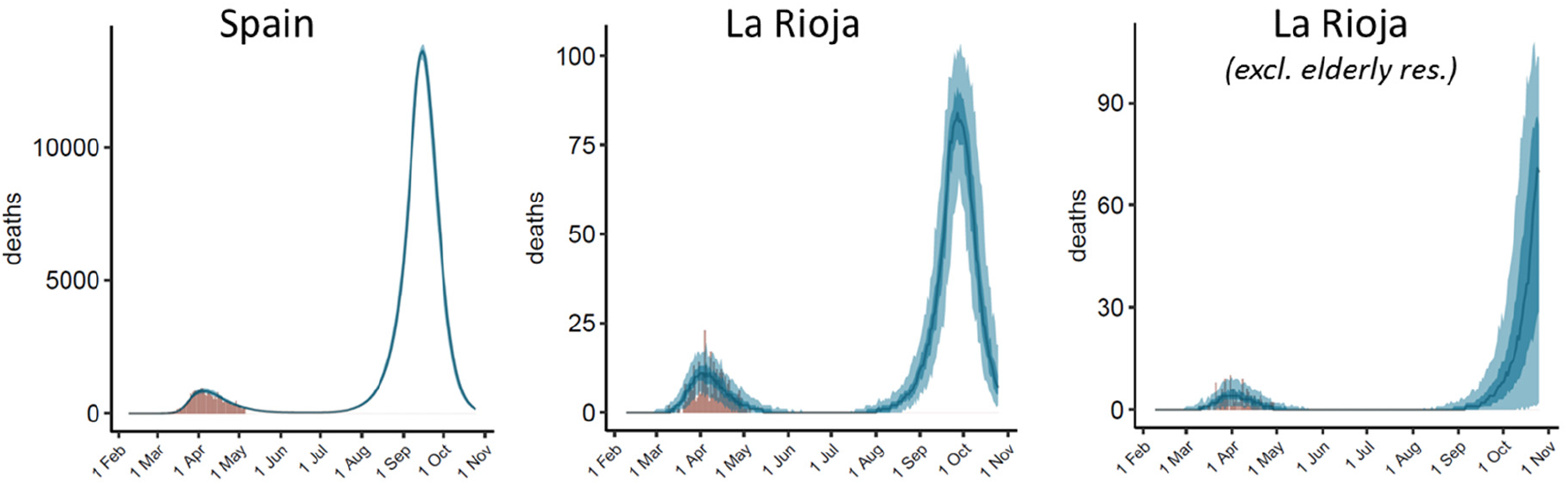
Estimated evolution of daily deaths over time in Spain and La Rioja (total data, and after excluding data from elderly residences) assuming *R_t_* = 1.0 between 11 May and 22 June 2020, and *R_t_* = 1.8 afterwards.

The model considers that all new infected cases have the same probability of infect new people (which is given by the SI distribution and the estimated *R*_t_ in eq. 1). However, in a situation as in La Rioja, where the cases inside the elderly retirement homes are reasonably isolated from the rest of the population, and outside the elderly residences the virtual totality of active cases seem to be detected and thus not likely to induce new infections, the effective *R*_t_ values in the analyzed dates could be much lower than those in the hypothetical scenarios discussed here, which would suggest a more optimistic situation for the upcoming months. In any case, after June 22^th^, with travels between provinces in Spain allowed, it would be essential to be able to detect any new focus of infection that might induce a sudden outbreak. For comparison, assuming the same scenario in Iceland (*R*_t_ = 1.0 between 11 May and 22 June 2020, and *R*_t_ = 1.8 afterwards), no outbreak is predicted in the studied period, even when assuming return to complete normality on 1 September 2020 (data not shown). The model can thus be useful for evaluating the long-term impact of the implementation and/or removal of intervention measures on the disease evolution in a given country or region.

### Limitations of the model and perspectives

The model used here depends strongly on the serial interval and infection-to-death probability distributions. Future updates of the model could benefit from the use of other available models for serial interval and infection to death distributions [16]. In some of these models, the incubation period averages around 5 days (CI 95% 2-14), with a median time delay of 13-17 days from illness onset to death, depending on the type of truncation [17]. In another study, using the most reliable data among their sets, the median serial interval was estimated at 4.6 days (95% CI: 3.5-5.9) [18]. A recently proposed model of SARS-CoV-2 transmission assumed a latent period of 4.6 days and an infectious period of 5 days, informed by the best-fit values for other betacoronaviruses [2]. Finally, a recent study estimated the median incubation period as 5.1 days (95% CI 4.5-5.8) [19]. Other parameter that can strongly affect the estimated number of infections is the IFR value. As above mentioned, there are several estimates for IFR based on available data [6-8] and on mathematical models [20].

A major limitation of current model is related to the fact that *R* is actually a dynamic parameter that may change over time and adopt different values in each of the communities forming a country. However, here I am assuming that *R* is constant along long periods of time, and only change upon a specific intervention. While this assumption seems to be adequate to estimate abrupt changes in *R* after major interventions, in many situations *R* can change over time in a more subtle way, for instance depending on local actions, such as disease transmission control in specific communities (e.g. elderly residences, hospitals), or on behaviours changing over time, like self-awareness of the population. This seems the case in Iceland, in which *R_0_* is smaller than in other countries, probably because of a better control of the first detected cases thanks to a higher detection rate. Inclusion of a more dynamic *R_t_* may lead to significant improvements of the model, but also to increased noise in the fitting process unless more data can be considered.

Regarding the size of data, while larger sets of data in entire countries show softer distribution curves of reported infections and deaths, there is also an implicit difficulty in describing the disease evolution with a single model, because the transmission dynamics in a country is usually formed by the disease evolution in different communities. This seems to be the case in Italy, in which the fitted model expects a single peak after a major intervention, while the distribution of reported deaths over time seem to have a shoulder after the main peak (Figure 3). This might indicate spreading from the initial focus to the rest of regions, which can largely affect the transmission dynamics in Italy [21]. While this geographical transmission is not explicitly considered in the model, its application can be a complement to other studies aiming to understand the evolution of the disease in time and space. For instance, our model estimates a total of 396 infections (95% CI: 153-819) in Spain between February 9 and 14, 2020. The model cannot distinguish whether these were local infections or externally acquired, but the number is consistent with recent studies on the spread of disease in Spain in mid-February, based on phylogenetic studies using SARS-Cov-2 whole-genome sequencing data, which estimated the origin of two SARS-Cov-2 clusters in Spain around February 14 and 18, 2020) [22].

## CONCLUSIONS

A Bayesian model of disease transmission inferred from reported deaths, previously developed by researchers at Imperial College London, has been applied here to 11 countries to estimate the impact of different intervention measures. The model has been updated, and have independently applied it to countries and regions with high number of reported cases, or high-incidence related to their population. The model indicates that major intervention measures have a similar impact on the disease transmission in most of these countries, and provides realistic estimates of the total number of infections, active cases and future outcome. The model can also be useful to help planning changes in the implementation of control measures, but the reliability of the long-term predictions may depend on the moment of the epidemic outbreak.

## AVAILABILITY

The data and model scripts are available upon request.

## Data Availability

The data and model scripts are available upon request.

## ACKNOWLEDGMENTS

This work was supported by grants BIO2016-79930-R from the Spanish “Programa Estatal I+D+i”, and 2019AEP095 from CSIC.

